# Coronary Artery Calcium Scans Powered by Artificial Intelligence Predicts Atrial Fibrillation Comparably to Cardiac Magnetic Resonance Imaging: The Multi-Ethnic Study of Atherosclerosis (MESA)

**DOI:** 10.1101/2024.01.04.24300746

**Authors:** Morteza Naghavi, Anthony P. Reeves, Kyle Atlas, Dong Li, Hamidreza Goodarzynejad, Chenyu Zhang, Thomas Atlas, Claudia Henschke, Matthew J. Budoff, David Yankelevitz

**Affiliations:** HeartLung.AI, 2450 Holcombe, Houston, TX, 77021; Department of Electrical and Computer Engineering, Cornell University, 616 Thurston Ave. Ithaca, NY 14853; The Lundquist Institute, 1124 W Carson St, Torrance, CA 90502; Tustin Teleradiology, 13422 Newport Ave Suite I, Tustin, CA 92780; Mount Sinai Hospital, 1468 Madison Ave, New York, NY 10029

**Keywords:** left atrial volume, coronary artery calcium, cardiac magnetic resonance imaging, atrial fibrillation, artificial intelligence

## Abstract

**Background:** Applying artificial intelligence to coronary artery calcium computed tomography scan (AI-CAC) provides more actionable information beyond the Agatston coronary artery calcium (CAC) score. We have recently shown that AI-CAC automated left atrial (LA) volumetry enabled prediction of atrial fibrillation (AF) in as early as one year. In this study we evaluated the performance of AI-CAC automated LA volumetry versus LA volume measured by human experts using cardiac magnetic resonance imaging (CMRI) for predicting AF, and compared them with CHARGE-AF risk score, Agatston score, and NT-proBNP (BNP).

**Methods:** We used 15-year outcome data from 3552 asymptomatic individuals (52.2% women, ages 45-84 years) who underwent both CAC scans and CMRI in the baseline examination (2000-2002) of the Multi-Ethnic Study of Atherosclerosis (MESA). AI-CAC took on average 21 seconds per scan. CMRI LA volume was previously measured by human experts. Data on BNP, CHARGE-AF risk score and the Agatston score were obtained from MESA.

**Results:** Over 15 years follow-up, 562 cases of AF accrued. The ROC AUC for AI-CAC versus CMRI and CHARGE-AF were not significantly different (AUC 0.807, 0.808, 0.800 respectively, p=0.60). The AUC for BNP (0.707) and Agatston score (0.694) were significantly lower than the rest (p<.0001). AI-CAC and CMRI significantly improved the continuous Net Reclassification Index (NRI) for prediction of AF when added to CHARGE-AF risk score (0.28, 0.31), BNP (0.43, 0.32), and Agatston score (0.69, 0.41) respectively (p for all<0.0001).

**Conclusion:** AI-CAC automated LA volumetry and CMRI LA volume measured by human experts similarly predicted incident AF over 15 years.

## Introduction

Left atrial (LA) size normalized by the body surface area has been studied extensively as a predictor of incident atrial fibrillation (AF), stroke and other adverse cardiovascular events^1,2,3,4,5,6,7,8,9^. However, this valuable biomarker has not been widely introduced to patient care for AF prediction and stroke prevention. Contrast-enhanced cardiac magnetic resonance imaging (CMRI) is the gold standard for measuring LA volume. However, CMRI is more time-consuming, has a higher cost, and is not as widely available as a non-contrast cardiac computed tomography (CT) scan obtained for coronary artery calcium (CAC) score. It would be of great clinical value if LA volume could be measured robustly in a CAC scan without additional radiation exposure to patients or using any contrast enhanced agent^10^. Such an add-on measurement can offer valuable insights into a patient’s cardiovascular risk beyond the CAC score.

We have developed an artificial intelligence-enabled tool that automatically and reliably estimates cardiac chambers volume in non-contrast cardiac CT scans obtained for CAC score. This tool (AI-CAC) is very rapid (takes on average 21 seconds per scan) and reports the volume of all four cardiac chambers as well as the left ventricular mass. We have recently applied the AI-CAC to CAC scans obtained in the baseline examination (years 2000-2002) of Multi-Ethnic Study of Atherosclerosis (MESA) and demonstrated its ability to predict AF in as early as one year^11^. The aim of the current study was to evaluate the performance of AI-CAC automated LA volumetry versus LA volume measured by human experts using CMRI for predicting AF. Furthermore, we compared their predictive value with that of CHARGE-AF risk score, Agatston score, and NT-proBNP (BNP).

## Methods

### Study population

The Multi-Ethnic Study of Atherosclerosis (MESA) is a prospective, population-based, observational cohort study of 6,814 men and women without clinical cardiovascular disease (CVD) at the time of recruitment. Six field centers in the United States participated in the study. As part of the initial evaluation (2000-2002), participants received a comprehensive medical history, clinic examination, and laboratory tests. Demographic information, medical history, and medication use at baseline were obtained by self-report. An ECG-gated non-contrast CT was performed at the baseline examination to measure coronary artery calcium (CAC). Non-CT scan covariates included BNP and variables used in calculating the CHARGE-AF Risk Score. Details on BNP assays measurements are described below under BNP Measurement. Covariates used in CHARGE-AF Score for our analyses are age, gender, ethnicity, height, weight, systolic blood pressure, diastolic blood pressure, current smoking, hypertension medication, diabetes, which were obtained as a part of MESA baseline exam 1 previously described^12^. Additionally, CHARGE-AF Risk Score includes myocardial infarction and heart failure which were by default absent in the asymptomatic MESA population at baseline exam 1.

### Outcomes

Participants were contacted by telephone every 9-12 months during follow-up and asked to report all new cardiovascular diagnoses. International Classification of Disease (ICD) codes were obtained. Incident AF was identified by ICD codes 427.3× (version 9) or I48.x (version 10) from inpatient stays and, for participants enrolled in fee-for-service Medicare, from Medicare claims for outpatient and provider services. For participant reports of heart failure, coronary heart disease, stroke, and CVD mortality, detailed medical records were obtained, and diagnoses were adjudicated by the MESA Morbidity and Mortality Committee. Additionally, BNP data was obtained from MESA core laboratory for MESA exam 1 participants. A detailed study design for MESA has been published elsewhere^12^. MESA participants have been followed since the year 2000. Incident AF has been identified through December 2018.

### The AI-CAC Tool for Automated Cardiac Chambers Volumetry

The AI-CAC tool referred to in this manuscript is called AutoChamber^TM^ (HeartLung.AI, Houston, TX), a deep learning model that used TotalSegmentator^13^ as the base input and was further developed to segment not only each of the four cardiac chambers; left atrium (LA), left ventricle (LV), right atrium (RA), and right ventricle (RV) but also ascending aorta, aortic root and valves, pulmonary arteries, and several other components which are not presented here. The AI-estimated LA volumetry is the focus of this manuscript. Figure 1 shows the segmentations of cardiac chambers in color. The base architecture of the TotalSegmentator model was trained on 1139 cases with 447 cases of coronary CT angiography (CCTA) using nnU-Net, a self-configuring method for deep learning-based biomedical image segmentation^14^. The initial input training data were matched non-contrast and contrast-enhanced ECG-gated cardiac CT scans with 1.5 mm slice thickness. Because the images were taken from the same patients in the same session, registration was done with good alignment. Following this transfer of segmentations, a nnU-Net deep learning tool was used for training the model. Additionally, iterative training was implemented whereby human supervisors corrected errors made by the model, and the corrected data were used to further train the model, leading to improved accuracy. To standardize the comparison in MESA, cardiac chambers were reported by gender and ethnicity adjusted by body surface area (BSA) using residual adjustment techniques. (BSA: 0.007184 × (height(m)0.725) × (weight(kg)0.425)). Additionally, an internal reference was developed based on the field of view size and the posterior height of thoracic vertebral bones. This measure would be used whenever BSA information is unavailable, however it was not an issue in MESA. AutoChamber™ AI was run on 6043 non-contrast CAC scans that consented to commercial data usage out of the 6814 scans available in MESA exam 1. Expert rules built in the AI-model excluded 125 cases due to missing slices in image reconstruction created by some of the electron beam CT scanners used in MESA baseline. These cases were random, and our investigations did not reveal any particular association with dependent or independent variables in our study (see Results).

**Figure 1.**
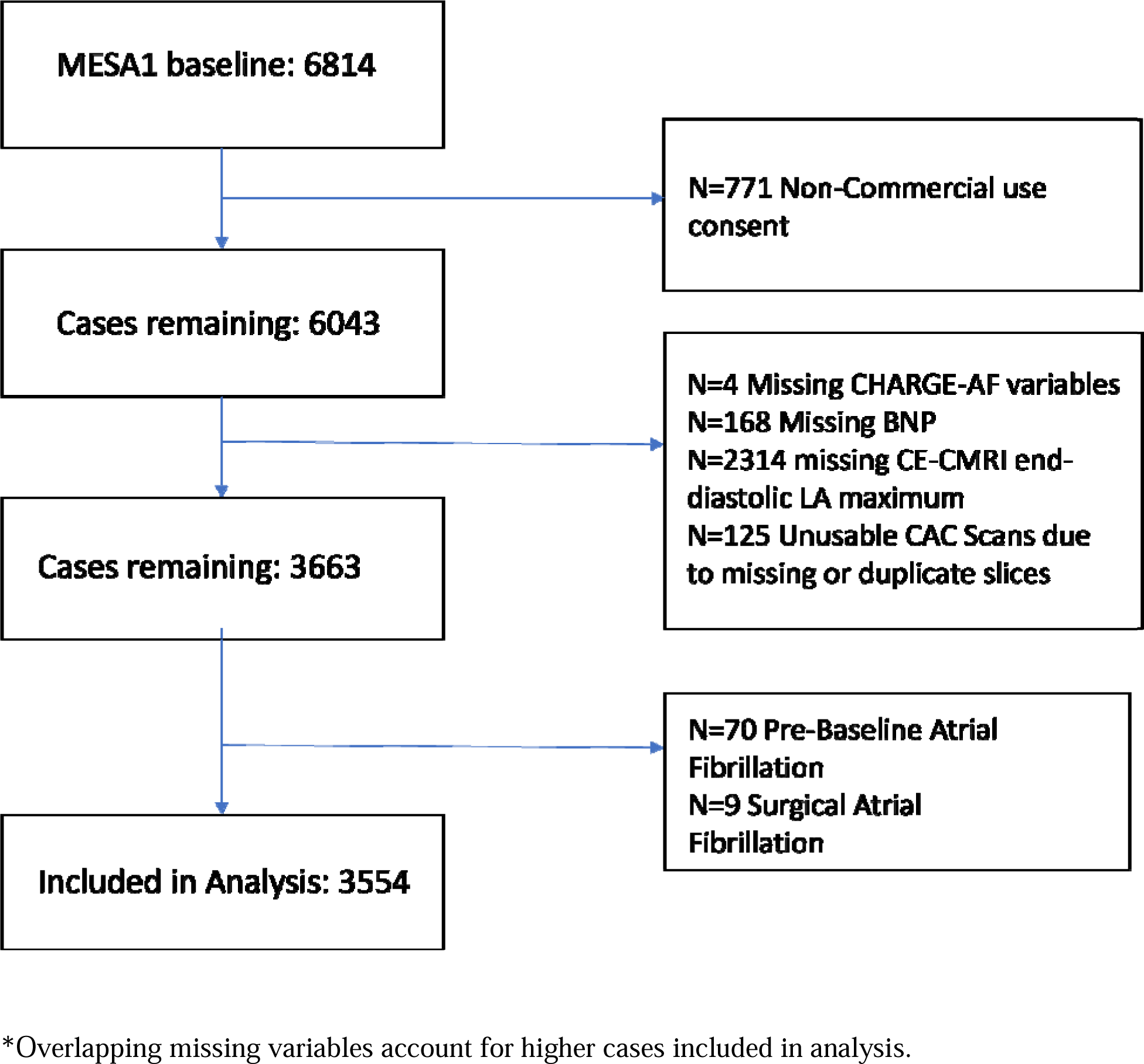
Flow chart depicting manuscript inclusion and exclusion criteria.

**Figure 2.**
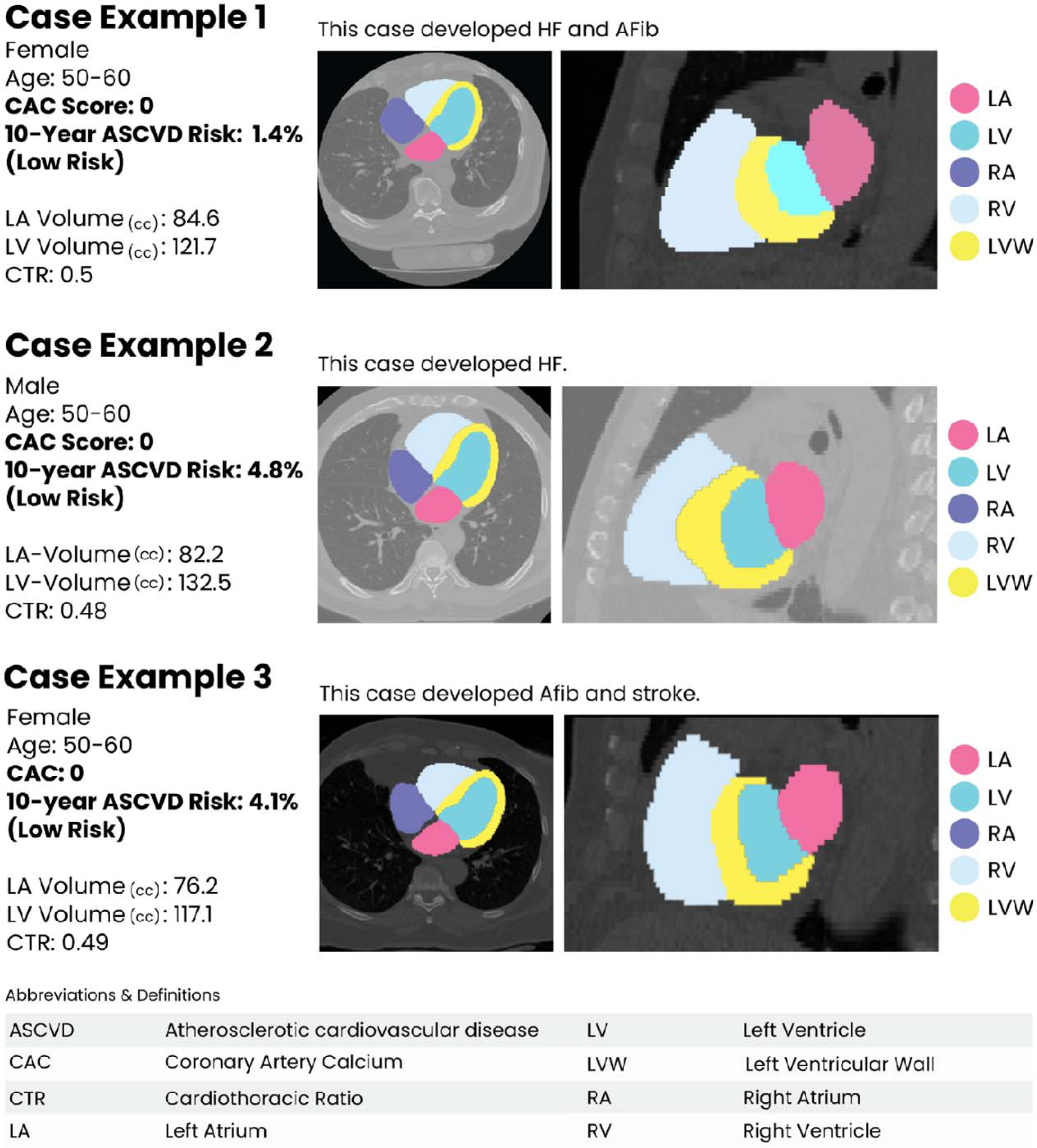
Examples of AI-CAC detection of high-risk individuals with enlarged left atrium (LA) in coronary artery calcium (CAC) scans with calcium score of zero who subsequently experienced adverse events.

### CMRI Measurement

CMRI is considered the most robust noninvasive imaging tool for studying cardiovascular structure and function. It creates detailed pictures of the beating heart and vessels to look at their anatomical and physiological properties. In MESA exam 1, CMRI was used to accurately quantify volume and dimensions of all four cardiac chambers during systole and diastole. For this study, the focus was end-diastolic LA maximum volume that was measured by mean transmit time (mL). CMRI can evaluate patterns of cardiac structural remodeling and the myocardial mechanical consequences of remodeling. Furthermore it can explore ventricular-arterial coupling, arterial stiffness, and other CMRI based imaging biomarkers^15^.

### CHARGE-AF Risk Score

The CHARGE-AF risk score was developed to predict risk of incident AF in three American cohorts, and it was validated in two European cohorts. The linear predictor from the CHARGE-AF Risk Score is calculated as: (age in years/5) * 0.5083+ethnicity (Caucasian/white) * 0.46491 + (height in centimeters/10) * 0.2478 + (weight in kg/15) * 0.1155 + (SBP in mm Hg/20) * 0.1972 – (DBP in mm Hg/10) * 0.1013+current smoking * 0.35931+antihypertensive medication use * 0.34889 + DM*0.23666 + heart failure(HF) * 0.701 + myocardial infarction(MI) * 0.496^16^. The result is the sum of the product of the regression coefficients and the predictor variables, which represents the change in the hazard ratio for a one-unit change in the corresponding predictor variable. Due to the asymptomatic cohort, presence of HF was not included in the equation.

### BNP Measurement

Details on BNP assays used in MESA have been reported^17^. N-terminal proBNP is more reproducible than BNP at the lower end of the distribution range, and more stable at room temperature. However, both BNP and N-terminal proBNP are clinically available. Intra-assay and inter-assay coefficients of variation at various concentrations of NT-proBNP have been previously reported^18,19^. The analytical measurement range for NT-proBNP in exam 1 was 4.9– 11699 pg/ml. The lower limits of detection for the NT-proBNP assay is 5 pg/mL, thus cases above 0 and below 4.99 were treated as 4.99 pg/mL. Clinically, values are not reported below 4.99 pg/mL because the analytical accuracy is poor at those low levels (i.e. typically a coefficient of variation of greater than 20% between repeat measures).

### Agatston CAC Score Measurement

Out of 6 study sites, three used cardiac-gated electron-beam computed tomography (CT) scans, whereas the other 3 sites used multidetector CT scans. Each participant was scanned twice at baseline examination, with mean Agatston score used for analysis^20^. All scans were phantom adjusted and read by 2 trained CT image analysts at a central MESA CT reading center, with high reproducibility and comparability between electron beam CT and multidetector CT scanning^21,22^. Detailed information on CT scan methods and interpretation has been given previously^21^.

CAC area and density were derived from total Agatston and volume scores, which were provided in the original MESA data set. The methods for this derivation are described in a previous article^23^.

### Statistical Analysis

We used SAS (SAS Institute Inc., Cary, NC) and Stata (StatCorp LLC, College Station, TX) software for statistical analyses. All values are reported as means ± SD except for BNP which did not show normal distribution and is presented in median and interquartile range (IQR). All tests of significance were two tailed, and significance was defined at the p<0.05 level.

Cumulative incidence was calculated using one minus the Kaplan-Meier survival estimate. Significance in cumulative incidence differences was determined using a test of proportions. The time-dependent ROC (receiver operator curve) AUC (area under the curve) was calculated using Cox proportional hazards regression at 15 years follow-up. Significance in AUC differences for the non-nested models were determined using the DeLong test.

BNP and CAC were natural logarithm-transformed (ln-transformed) to avoid undue influence of large values. CMRI LA end-diastolic maximum volume was used in all models. Category-free (continuous) net reclassification index (NRI) was calculated using the sum of the differences between the proportions of upward reclassifications and downward reclassifications for AF events and AF non-events, respectively. NRI was developed as a statistical measure to evaluate the improvement in risk prediction models when additional variables are incorporated into a base model^24^. We have analyzed data for AF prediction at 15 years follow-up.

### Ethical Approval

This study has received proper ethical oversight. All subjects gave their informed consent for inclusion before they participated in the study. Subjects who did not consent were removed from the study.

## Results

For our study, we removed 771 MESA participants who did not consent for commercial use of data, leaving 6043 participants for our analysis. After removing 125 cases with missing slices in CAC scans, 4 cases with missing data for CHARGE-AF Risk Score, and 168 cases with missing BNP values we have 5746 remaining participants. Subsequently, we have removed 70 cases with pre-baseline AF, 9 cases with surgical AF leaving 5567 cases. CMRI measurements for one or more variables were missing from 2430 participants, leaving 3552 cases available from analysis (Figure 1).

28.1% of participants were aged 45-54 years, 27.3% aged 55-64 years, 29.8% aged 65-74 years, and 14.8% aged 75-84 years.52.2% were women, 39.7% were White, 26.1% Black, 22% Hispanic, and 12.1% Chinese. Table 1 shows the baseline characteristics of MESA participants who were diagnosed with incident AF versus those who were not over the period of 15 years follow up. Over 15 years follow up 562 cases of AF accrued. In univariate comparisons, incident AF cases were older, more likely male, and more likely White. The incident AF cases for AI-CAC had higher cardiac chamber volumes for LA, LV, RA, LV Wall, and CMRI had higher cardiac chamber volumes for LA, LV, and LV Wall. CHARGE-AF Risk Scores, Agatston CAC score, and NT-proBNP levels were elevated in incident AF cases versus those without incident AF (all comparisons p< 0.001) (Table 1).

**Table 1:**
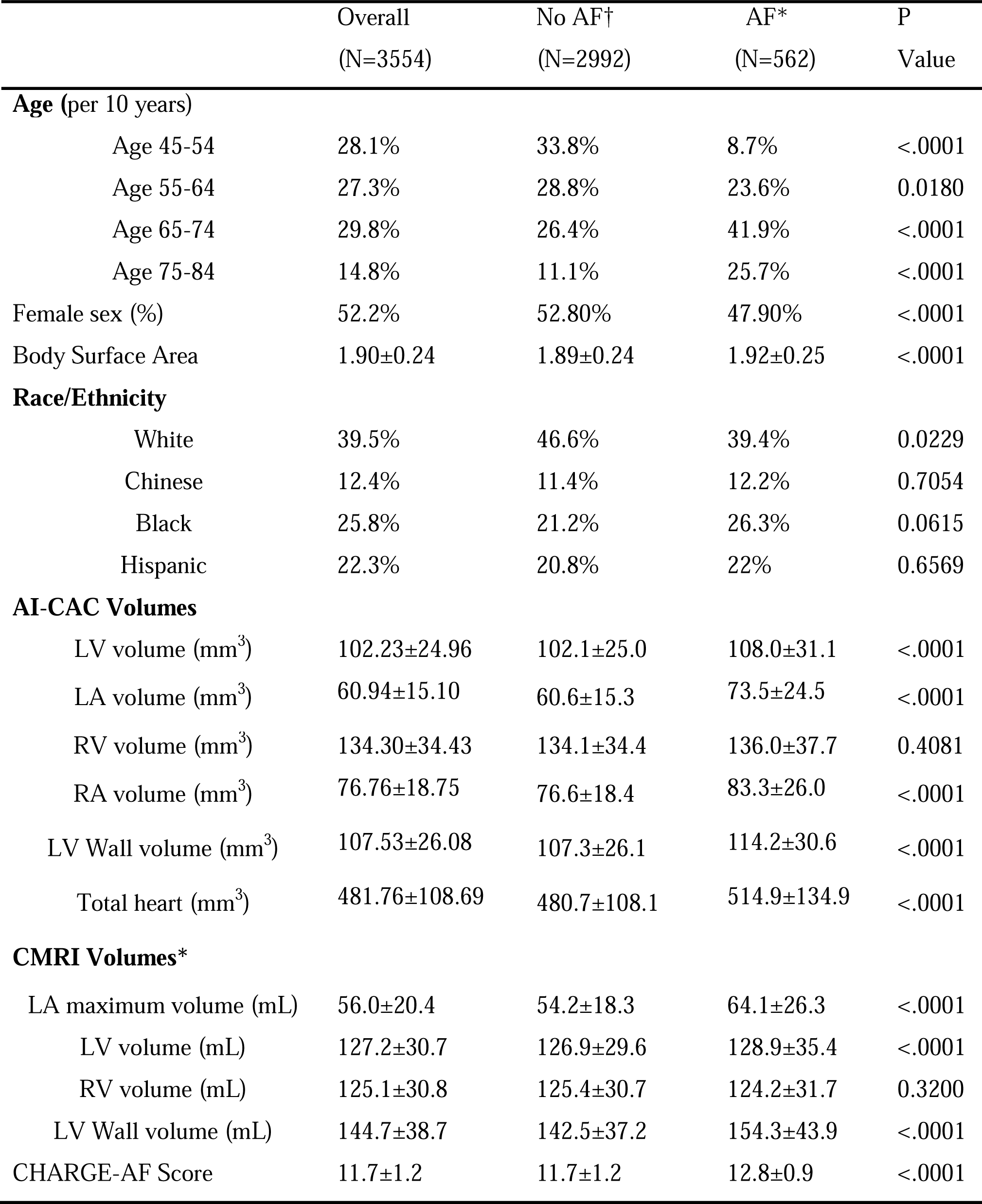
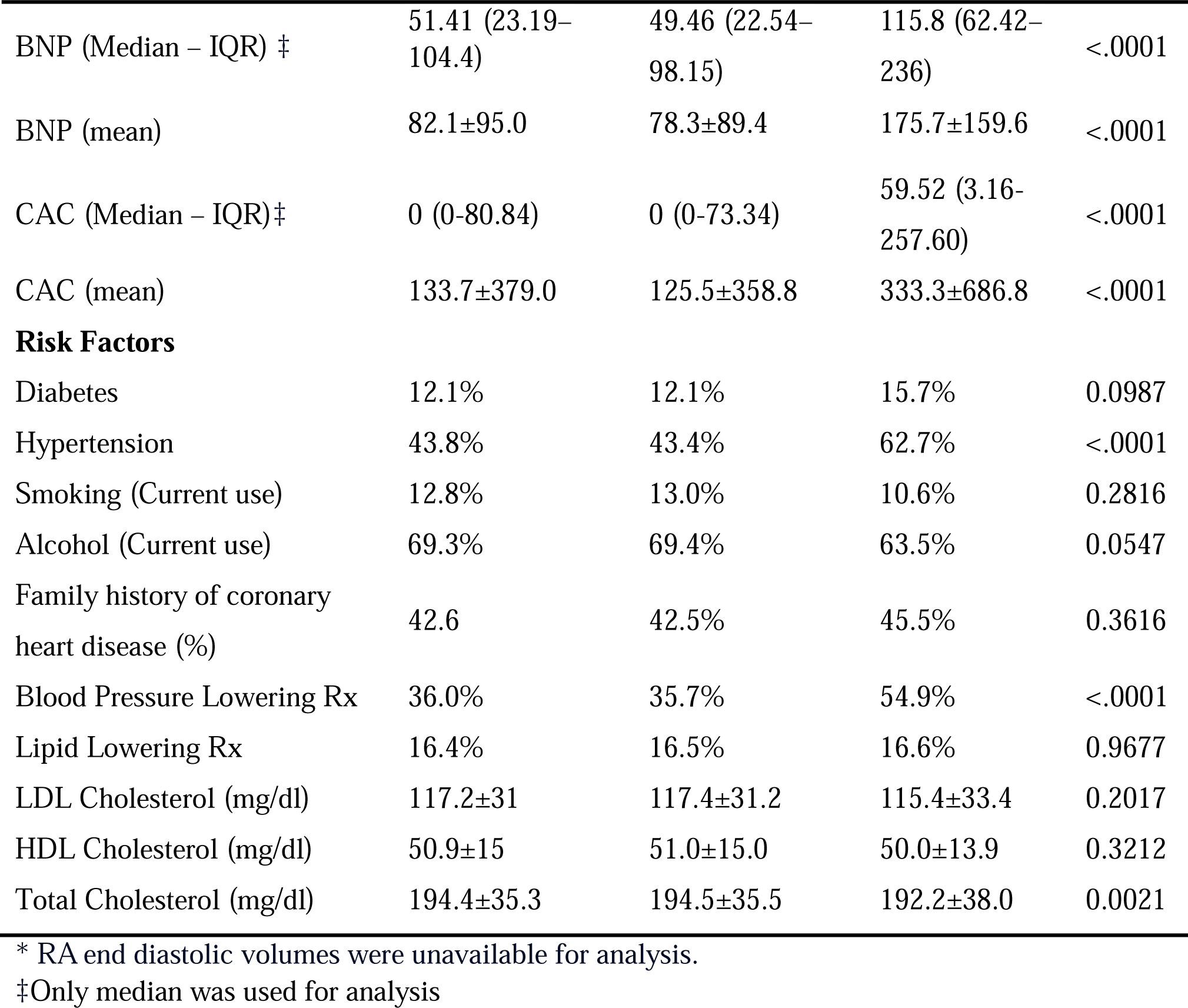
Baseline characteristics of the Multi-Ethnic Study of Atherosclerosis (MESA) participants including cases with and without Atrial Fibrillation (AF) at 15 years.

The cumulative incidence of AF over 15 years for AI-estimated LA volume, CMRI estimated LA volume, CHARGE-AF Risk Score, NT-proBNP and CAC were not significantly different (Figure 3a-e). The incidence of AF in the 95^th^ percentile of AI-LA volume, CMRI LA volume, Agatston CAC score, CHARGE-AF Risk Score, and BNP were 45.2%, 37.9%, 46.4%, 49.8%, and 45.5%, respectively.

**Figure 3a-e.**
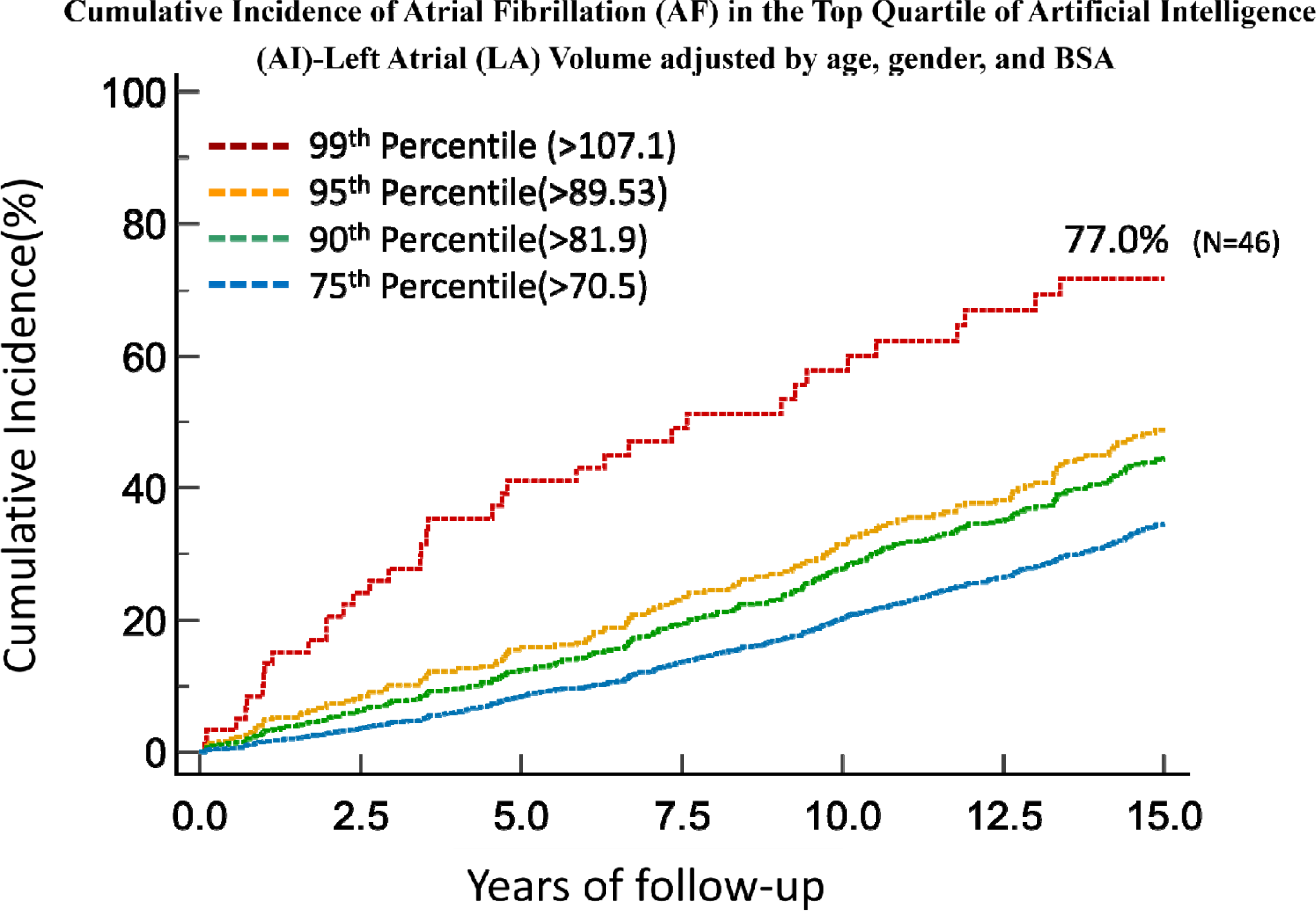
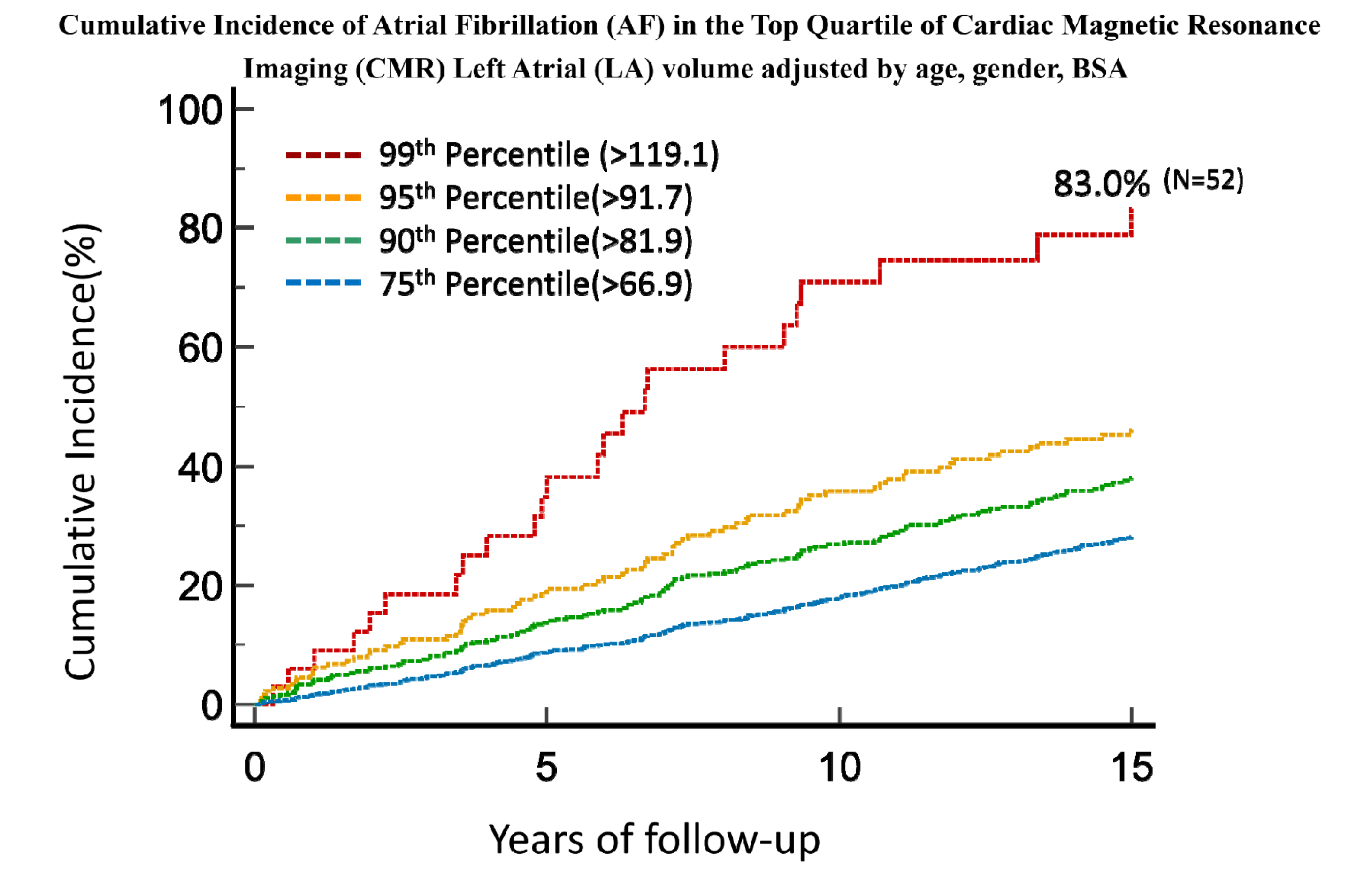
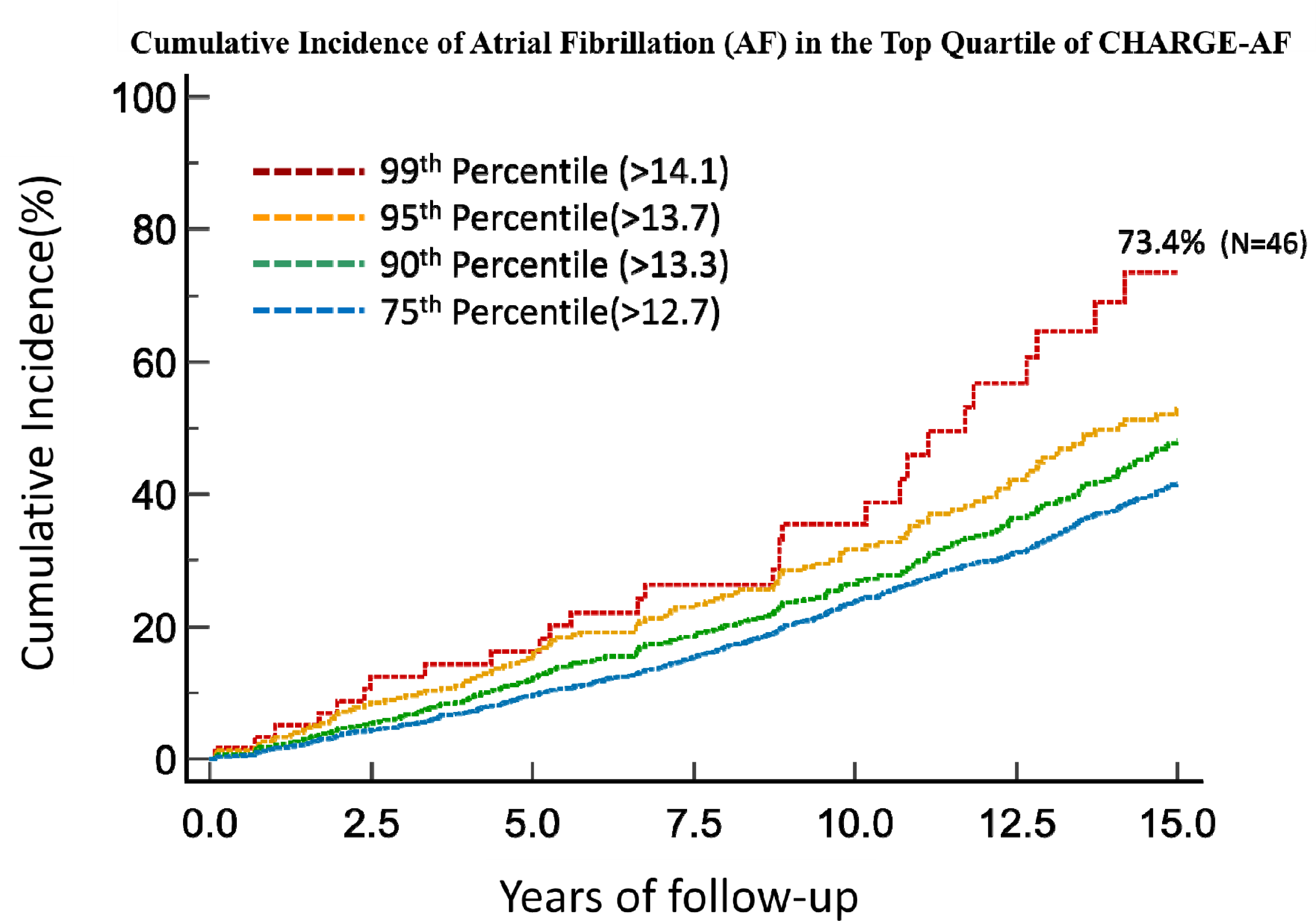
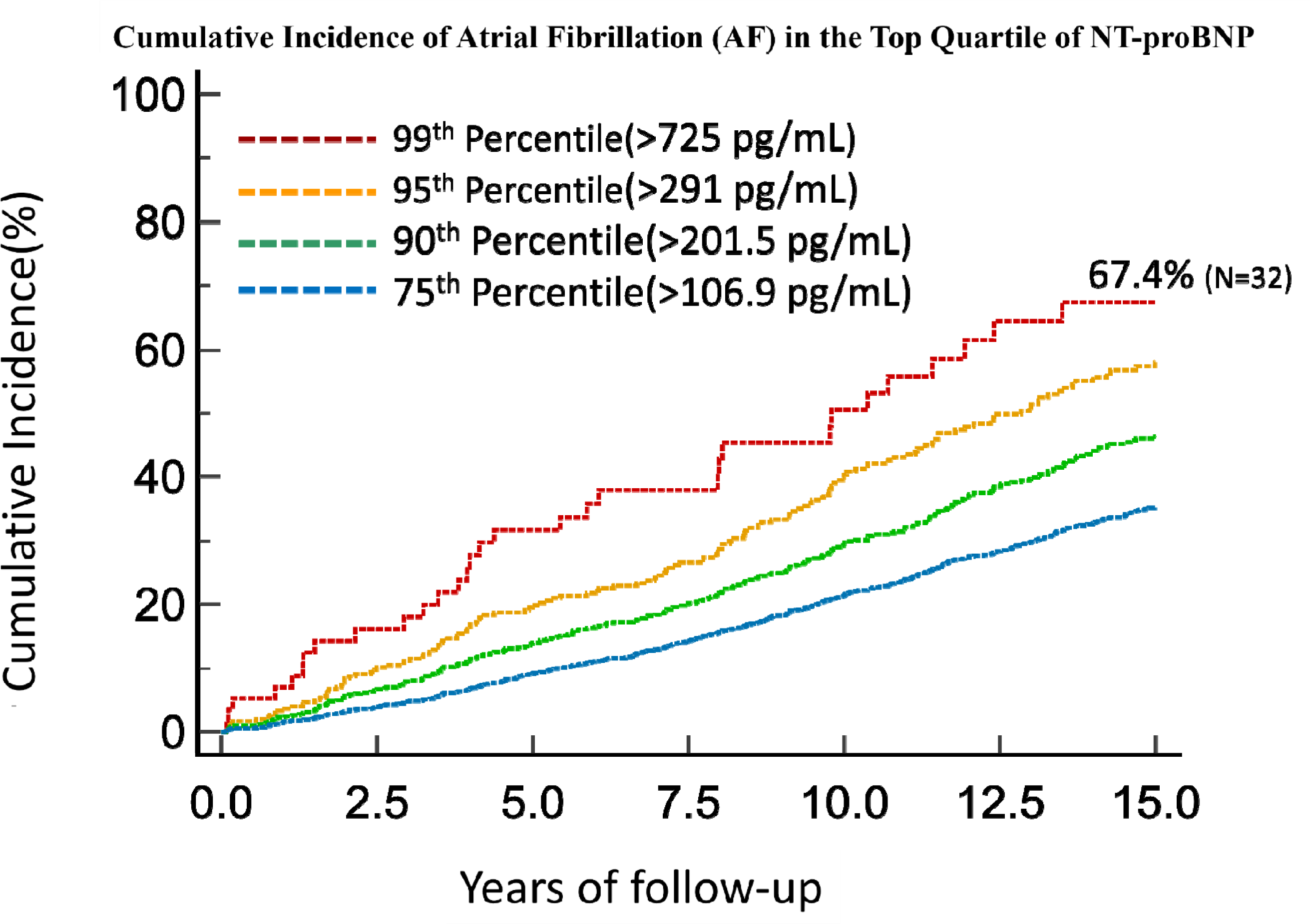
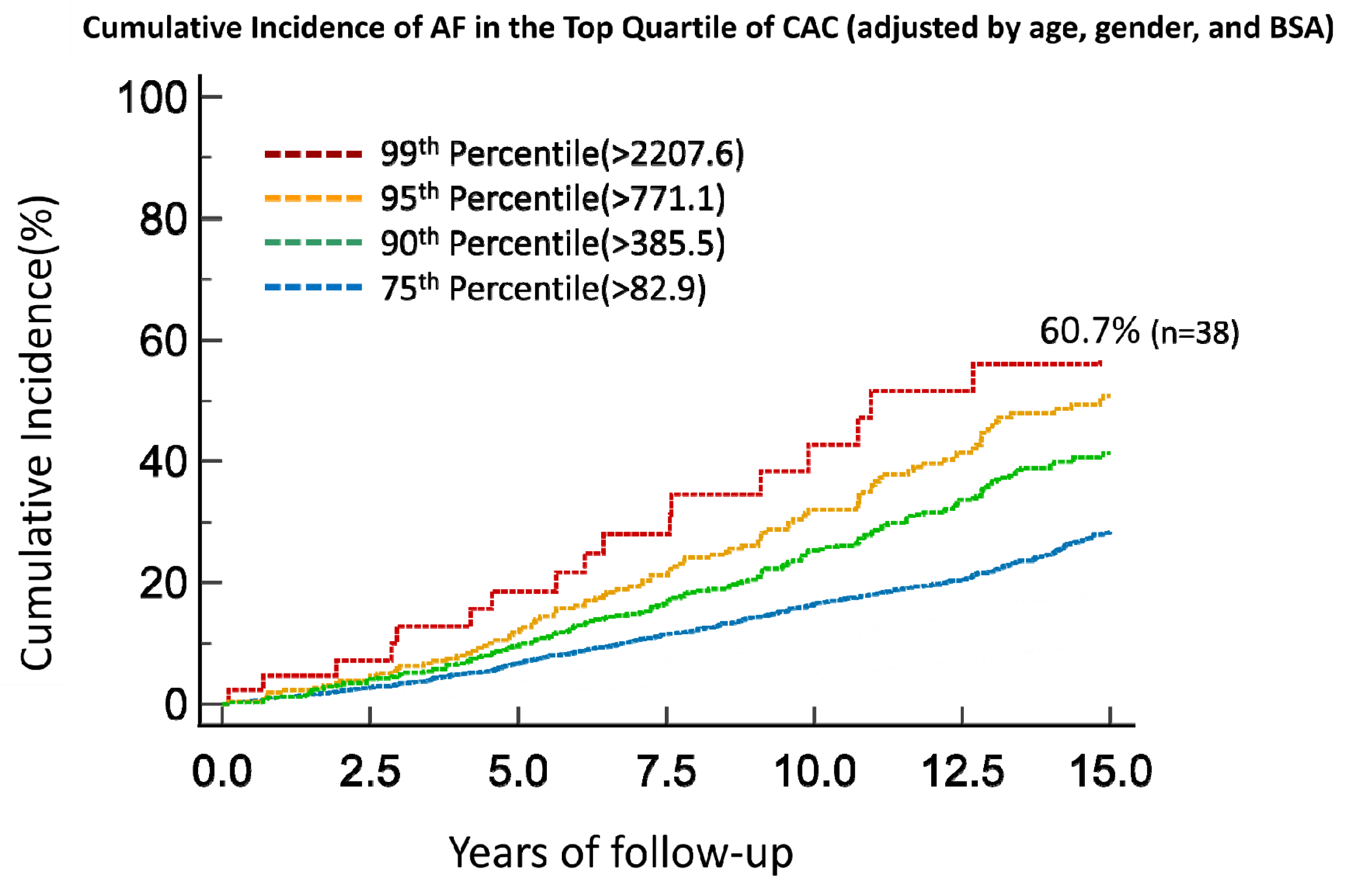
Cumulative Incidence of Atrial Fibrillation (AF) in the Top Quartile of AI-CAC Left Atrial (LA) Volume, CHARGE-AF Score, NT-proBNP (BNP), Agatston CAC Score, and Cardiac Magnetic Resonance Imaging (CMRI) LA Volume over 15 years of follow-up in the Multi-ethnic Study of Atherosclerosis (MESA).

The AUC for 15-year AF prediction by AI-estimated LA volume (adjusted by age, gender, BSA) was significantly higher than AUC for Agatston CAC Score and BNP (p<.0001), and comparable to CHARGE-AF and CMRI LA volume (Figure 4).

**Figure 4.**
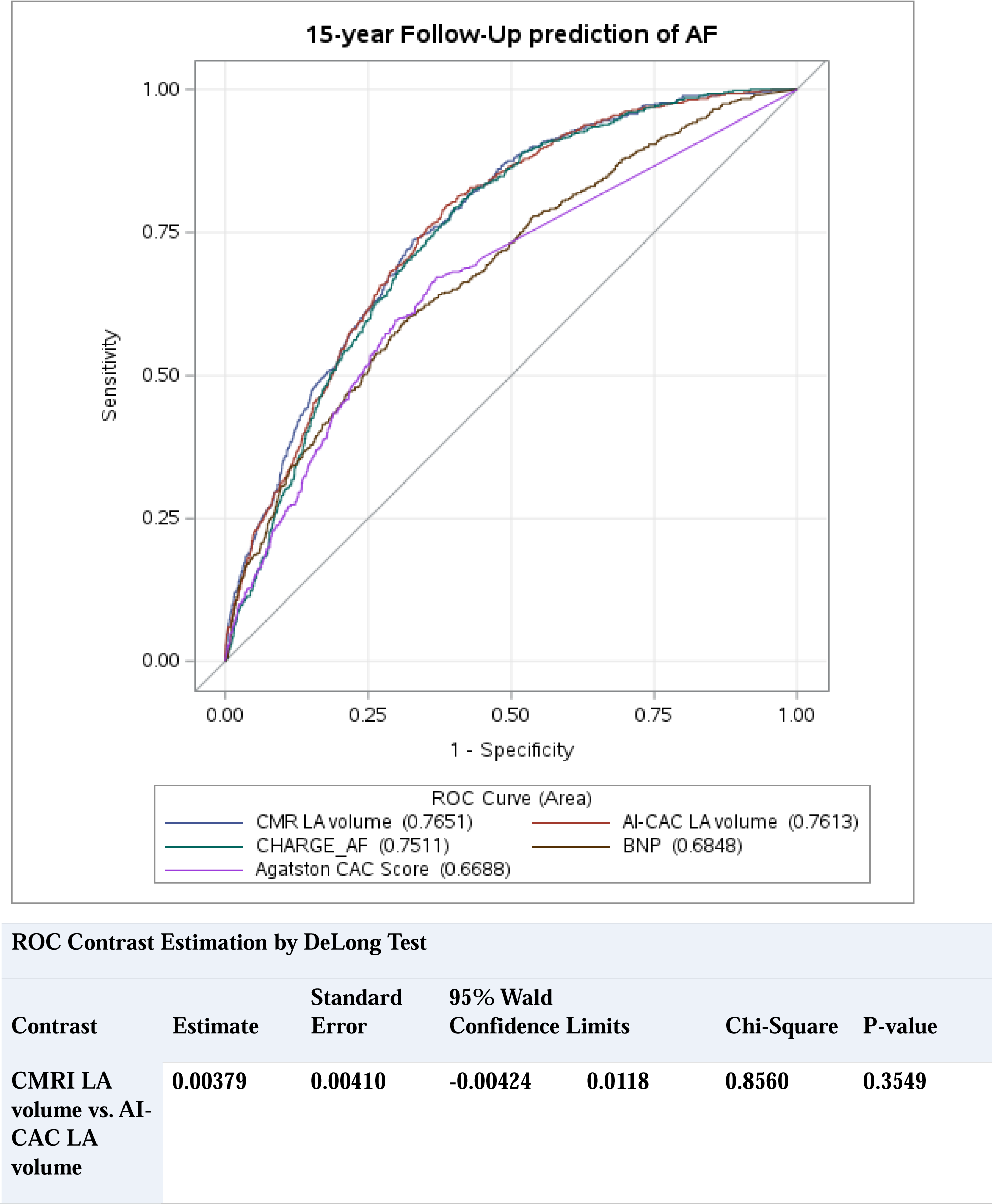
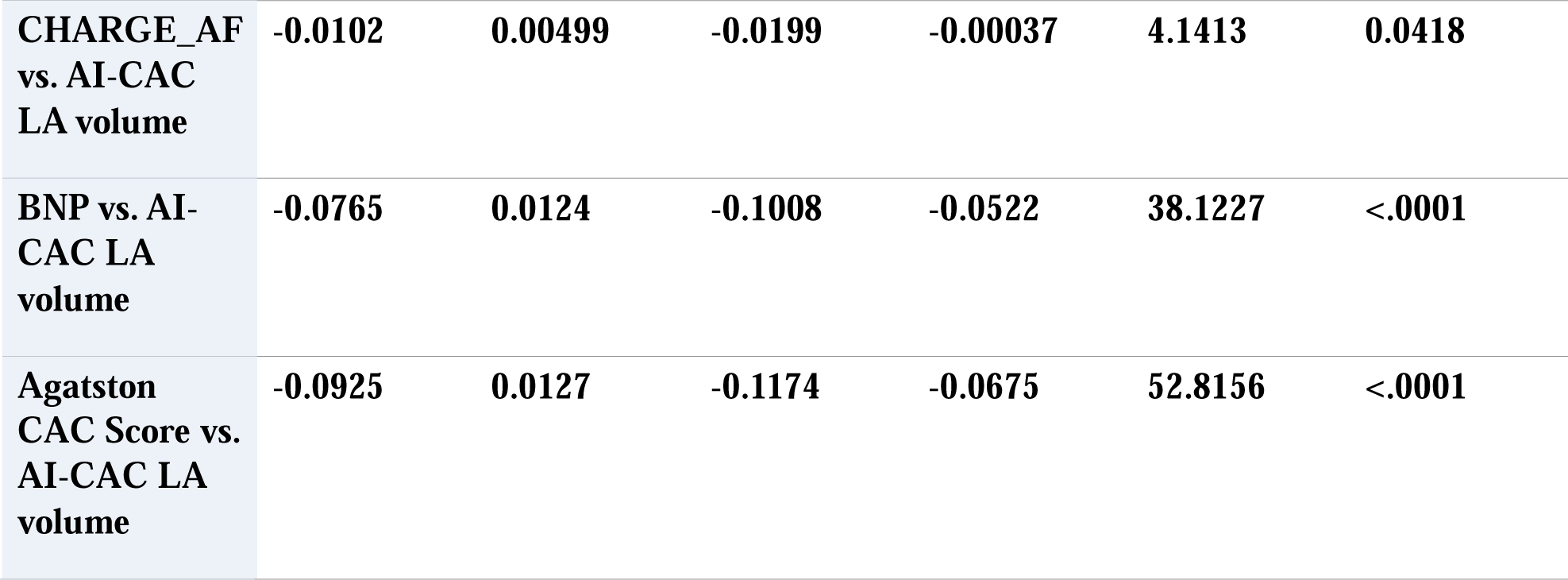
Comparing Receiver Operating Curve (ROC) Area Under Curve (AUC) between AI-CAC Left Atrial (LA) Volume Alone vs NT-proBNP (BNP), CHARGE-AF, and Cardiac Magnetic Resonance Imaging (CMRI) LA Volume.

The continuous NRI for prediction of AF when AI-estimated LA volume and CMRI LA volume was added to CAC score as the only predictor in the base model was highly significant (0.69, 0.41, respectively p<0.0001). Similarly, the NRI for AI-LA volume and CMRI LA volume when added to base model with CHARGE-AF Risk Score (0.28, 0.31) and BNP (0.43, 0.32), respectively, was significant (p<.0001). NRI was not calculated for AI-CAC LA volume with CMRI LA volume.

A significant number of low-risk participants with CAC 0 have enlarged cardiac chambers (Figure 2). Examples of three high risk patients with enlarged LA and LV volume with CAC score 0 and CAC below 50^th^ percentile, who are currently categorized as low risk have been provided. A significant number of high-risk patients with enlarged LA volume but CAC score 0 and CAC below 50^th^ percentile are currently categorized as low risk (Figure 5).

**Figure 5.**
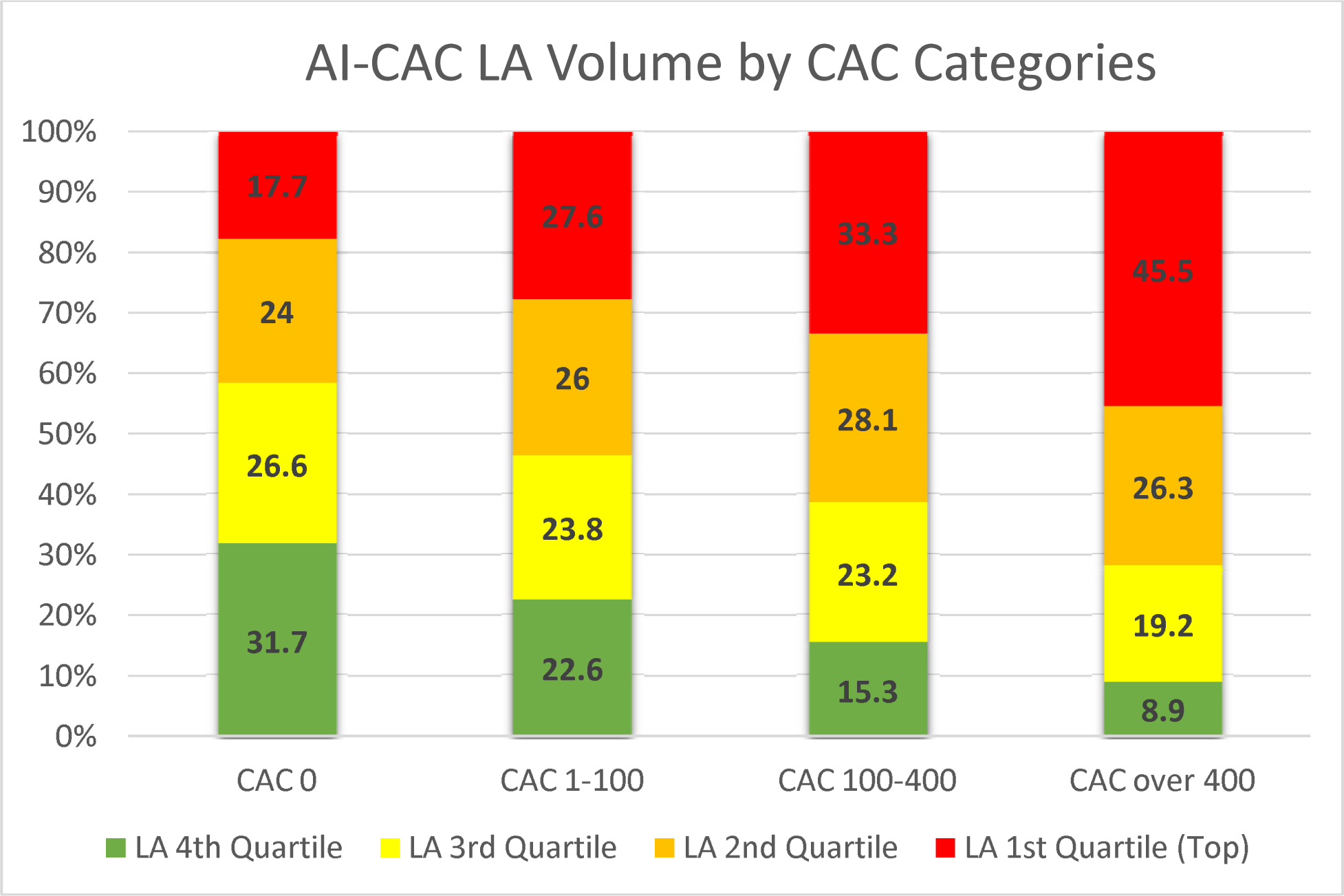
Quartiles of AI-CAC Left Atrial (LA) Volume by Coronary Artery Calcium (CAC) Score Categories.

The 125 cases with missing slices were 49.8% male and 50.2% female. None of these cases had a diagnosis of AF. As there were no associations with dependent or independent variables in our study, losses were likely random.

## Discussion

To our knowledge this is the first report comparing the predictive value of LA volume for predicting AF measured by an AI tool in non-contrast cardiac CT scans versus human experts in contrast-enhanced cardiac MRI. Our study demonstrated that LA volumetry using AI-CAC has comparable predictive value to that of the gold standard (CMRI). Both AI-CAC and CMRI provided for a sizable net reclassification improvement on top of CHARGE-AF risk score, BNP, and Agatston score. The moderately strong correlation between AI-CAC and CMRI LA volume (R=0.62) is evident despite the known asynchrony between these two measurements with cardiac CT measurements being done at around 70-80% end-diastole, versus CMRI at about 100% end-diastole.

AF is the most common sustained cardiac arrhythmia with an increased risk of cardiovascular disease outcomes including a five-fold higher risk of stroke^25^. However, it often goes undiagnosed in approximately 30% of cases due to its paroxysmal nature and minimal or even absent symptoms^26^. In the United States alone, it is estimated that around one million cases of AF are asymptomatic or undiagnosed^27^. Consequently, while the risk of stroke in AF patients can be reduced by two-thirds with oral anticoagulation therapy, more than 10% of patients experience their first manifestation of the undiagnosed AF as a stroke. A reasonable intervention following the detection of individuals at high risk of AF would be recommending a warble ECG monitor to watch for episodes of asymptomatic AF.

Clinical risk assessment tools might identify individuals for targeted screening of AF. A recent meta-analysis identified fourteen established risk prediction models and one of the best performing models showed to be CHARGE-AF. Sinner et al showed adding BNP, substantially improved AF risk prediction beyond clinical factors^28^. Therefore, a combination of CHARGE-AF and BNP can be a significant player in AF prediction. However, individualized assessment of LA size can be a more direct and specific indicator of risk. Among the various measurements of left atrial size, maximal left atrial volume indexed to body surface area stands out as the most robustly linked to cardiovascular conditions; it exhibits the highest sensitivity in forecasting cardiovascular events and offers consistent and precise risk assessment^1^.

### AI-powered CAC Scans

Since 1990 when Agatston and Janowitz 19 introduced their coronary artery calcium scoring technique, there has been little to no advancement in CAC scans and CAC scoring. Despite significant advancements in cardiac CT imaging and the transition from electron beam CT to multi-detector CT, patients today receive the report from CAC that was given some 20 years ago. In the meantime, numerous research studies have shown the value of non-coronary findings in CAC scans.^6,7,8,9,22,29^. Our study brings to light the practical use of non-coronary findings in CAC scans and corroborates recommendations by Heinz Nixdorf Recall Study investigators for a comprehensive CVD risk assessment in CAC scans beyond the CAC score and coronary heart disease^6,7,8,9^. It is noteworthy that the application of AI enabled cardiac chambers volumetry is not limited to CAC scans. In fact, it can be replicated in non-gated lung cancer screening scans. We have recently demonstrated in 169 patients with ECG-gated cardiac and non-gated lung CT scans in the same patients (paired scans done same day) that cardiac chambers volume measure in the two scans were strongly correlated (R^2^= 0.85-0.95 for different chambers, all p values <0.001)^30^.

Our study has some limitations. The CHARGE-AF Risk Score includes presence of HF and MI, but due to the asymptomatic nature of MESA these variables were excluded. The MESA Exam 1 baseline CT scans, performed between 2000 and 2002, were predominantly conducted using electron-beam computed tomography (EBCT) scanners. This technology is no longer the commonly used method of CAC scanning. Since our AI training was done completely outside of MESA and used a modern multi-detector (256 slice) scanner, we do not anticipate this to affect the generalizability of our findings. Because MESA used the ICD codes to identify a history of AF at baseline and newly diagnosed AF, and it is known that ICD based diagnosis can be inaccurate (PPV 70–96%, median sensitivity 79%)^31^ it is likely that MESA missed some cases of AF.

## Conclusion

AI-CAC automated LA volumetry predicted AF as strongly as LA volume measured by human experts in CMRI. Both AI-CAC and CMRI outperformed BNP and the Agatston score for AF prediction. The clinical utility of AI-enabled automated cardiac chambers volumetry as an added value to CAC scans is significant and warrants further investigations.

## Data Availability

All data is obtained from MESA Exam 1 CT and CMRI.

## Funding and Acknowledgement

This research was supported by 2R42AR070713 and R01HL146666 and MESA was supported by contracts 75N92020D00001, HHSN268201500003I, N01-HC-95159, 75N92020D00005, N01-HC-95160, 75N92020D00002, N01-HC-95161, 75N92020D00003, N01-HC-95162, 75N92020D00006, N01-HC-95163, 75N92020D00004, N01-HC-95164, 75N92020D00007, N01-HC-95165, N01-HC-95166, N01-HC-95167, N01-HC-95168 and N01-HC-95169 from the National Heart, Lung, and Blood Institute, and by grants UL1-TR-000040, UL1-TR-001079, and UL1-TR-001420 from the National Center for Advancing Translational Sciences (NCATS).

## Acknowledgement

This research was supported by 2R42AR070713 and R01HL146666 and MESA was supported by contracts 75N92020D00001, HHSN268201500003I, N01-HC-95159, 75N92020D00005, N01-HC-95160, 75N92020D00002, N01-HC-95161, 75N92020D00003, N01-HC-95162, 75N92020D00006, N01-HC-95163, 75N92020D00004, N01-HC-95164, 75N92020D00007, N01-HC-95165, N01-HC-95166, N01-HC-95167, N01-HC-95168 and N01-HC-95169 from the National Heart, Lung, and Blood Institute, and by grants UL1-TR-000040, UL1-TR-001079, and UL1-TR-001420 from the National Center for Advancing Translational Sciences (NCATS). The authors thank the other investigators, the staff, and the participants of the MESA study for their valuable contributions. A full list of participating MESA investigators and institutions can be found at http://www.mesa-nhlbi.org

